# ASYMPTOMATIC MONKEY POX VIRUS INFECTION: A SELF-SAMPLING SCREENING INTERVENTION ADRESSED TO GAY, BISEXUAL AND OTHER MEN WHO HAVE SEX WITH MEN AND TRANS WOMEN IN SPAIN

**DOI:** 10.1101/2023.02.20.23286168

**Authors:** Cristina Agustí, Héctor Martínez-Riveros, Àgueda Hernández-Rodríguez, Cristina Casañ, Yesika Díaz, Lucía Alonso, Elisa Martró, Jordana Muñoz-Basagoiti, Marçal Gallemí, Cinta Folch, Ibrahim Sönmez, Héctor Adell, Marta Villar, Alexia París de León, Sandra Martinez-Puchol, A.C Pelegrin, Daniel Perez-Zsolt, Dàlia Raïch-Regué, Rubén Mora, Luis Villegas, Bonaventura Clotet, Nuria Izquierdo-Useros, Pere-Joan Cardona, Jordi Casabona

## Abstract

We aimed to assess the prevalence of asymptomatic cases of monkeypox virus (MPXV) infection among gay, bisexual, and other men who have sex with men and trans women (TW), using a self-sampling strategy. Anal and pharyngeal swabs were tested by MPXV real-time PCR and positive samples inoculated into Vero E6 cells, which were subsequently checked for cytopathic effect (CPE).

Seven out 113 participants were MPXV positive (6.19% (95% CI: 1.75%-10.64%)). Five tested positive in pharyngeal swabs, one in anal swab and one in both. Six did not present symptoms recognized as MPXV infection. Three samples were positive for CPE, and showed anti-vaccinia pAb staining by FACS and confocal microscopy.

We describe Mpox cases that remain undiagnosed and show reproductive virus despite low viral loads and who might be able to infect others. Restricting testing to individuals reporting Mpox symptoms may not be enough to contain outbreaks.

## Introduction

Mpox is a zoonotic disease caused by monkeypox virus (MPXV), a virus belonging to the Orthopoxvirus genus, which is endemic in several African countries^1^. From 1 January through 12 December 2022, a cumulative total of 82,628 laboratory-confirmed cases of Mpox and 65 deaths were reported to the World Health Organization (WHO) from 110 countries^2^. On 23 July 2022 the WHO declared Mpox to be a Public Health Emergency of International Concern^3^. Spain, with 7,412 cases, has been the third-most affected country after the United States of America and Brazil^2^. In Spain, as in other countries, the outbreak has mainly affected gay, bisexual and other men who have sex with men (GBMSM) with no documented history of travel to countries where MPXV is endemic.

MPXV infection can cause genital, perianal, oral lesions as well as complications like proctitis and tonsillitis^4^. Although, sexual transmission by means of semen has not been ruled out^5^, some authors suggest that rather than the respiratory route, local inoculation by close skin-to-skin contact during sexual activity is the dominant transmissibility mode of MPXV, in non-endemic Mpox countries^6^.

Diagnosis of MPXV infection is based on nucleic acid amplification testing, using quantitative or conventional polymerase chain reaction (PCR). The recommended specimen type for laboratory confirmation of MPXV is skin lesion material including; swabs of lesion surface and/or exudate, roofs from more than one lesion, or lesion crusts^7^. MPXV testing is recommended for suspected cases presenting with symptoms that suggest this type of infection. However, two previous studies have reported positive MPXV PCR results among asymptomatic individuals^8, 9^. This supports the hypothesis that a proportion of MPXV infections remain undiagnosed, either because individuals have no symptoms (asymptomatic/pre-symptomatic infections), or because their symptoms are not attributed to a possible MPXV infection (unrecognized infections)^9^. Furthermore, very few studies have explored whether these infections could contribute to viral transmission between individuals.

Ward et al.^10^ performed a contact tracing study, linking data on case-contact pairs of MPXV infection on probable exposure dates in the UK between 6 May and 1 August 2022. They demonstrated that more than half (53%) of the transmission events in the UK outbreak occurred in the pre-symptomatic phase of infection. Furthermore, they estimated that transmission occurred up to four days before the onset of symptoms^10^. Retrospective PCR detection in patients of sexual health clinics in France and Belgium suggests that some patients could have an asymptomatic MPXV infection^8, 9^. In symptomatic Mpox cases, a study performed in a cohort of patients with relatively mild disease showed MPXV viral clearance within the first 2 months following the appearance of symptoms and a shorter duration of the period when replication-competent virus was detected in viral cultures^11^

The transmission dynamics of MPXV in the current outbreak are highly consistent with a sexually transmitted infection (STI)^12^. Although a few women and children have been infected since May 2022, the majority of cases in Spain have occurred among GBMSM. The 2022 MPXV outbreak shows some similarities with the HIV epidemic regarding potential stigmatization of key populations, such as GBMSM^13^. Stigma can prevent access to care for diagnosis and, in turn, prevent contact tracing and other containment measures. Alternative STI testing modalities such as self-testing and self-sampling constitute important options to diversify and optimize testing access and studies have demonstrated that they increase uptake of STI testing for all groups, including those at high-risk ^14–16^. Furthermore, a recent study showed that the performance of diagnostic tests from self-collected samples was similar to that of physician-collected samples, suggesting that self-sampling is a reliable strategy for diagnosing MPXV infection^17^.

In the present study, we aimed (i) to assess the prevalence of MPXV infection among asymptomatic highly exposed GBMSM and trans women (TW) who were recruited in a community-based centre in Barcelona, (ii) to assess the potential transmissibility of MPXV and (iii) to evaluate the feasibility and acceptability of a community-based self-sampling strategy for Mpox diagnosis.

## Results

### Characteristics of participants

From August to October 2022, 113 individuals participated in the study. The main characteristics of the participants are shown in **Table 1**. From all the participants, 89 (78.76%) were cis men, 17 (15.04%) were TW and 3 (2.65%) non-binary gender. The median age of participants was 35.0 years (Interquartile Range (IQR): 30.0-43.0), 96 (85.02%) individuals were gay or bisexual and 72 (63.72%) were migrants Additionally, 44 (38.94%) participants self-reported HIV infection and among HIV negative participants 41 (59.42%) were on PreP and 58 (51.33%) had had an STI in the previous 12 months. Regarding MPXV, 28 (24.78%) participants had had contact with a confirmed Mpox case over the previous 30 days. Also 7 (6.19%) and 13 (11.50%) participants had received the Mpox vaccine in their childhood or in the previous 12 months, respectively. In addition, 80 (70.80%) individuals were extremely or moderately concerned about Mpox and 53 (46.90%) considered it likely or very likely that they would get an MPXV infection (**Table 1**).

**Table 1.** Main characteristics of participants from Stop Mpox. August - October 2022. Barcelona (Spain). N: 113

|  | Monkeypox virus infection status |  |  | p value |
| --- | --- | --- | --- | --- |
|  | All N=113 | Negative N=106 | Positive N=7 |  |
| **Median age (IQR)** | 35.00<br>[30.00;43.00] | 35.00<br>[30.00;43.00] | 37.00<br>[34.00;48.50] | 0.235 |
| **Gender** |  |  |  | 0.744 |
| Cis man | 89 (78.76%) | 82 (77.36%) | 7 (100.00%) |  |
| Trans woman | 17 (15.04%) | 17 (16.04%) | 0 (0.00%) |  |
| Non binary person | 3 (2.65%) | 3 (2.83%) | 0 (0.00%) |  |
| DK/DA | 4 (3.54%) | 4 (3.77%) | 0 (0.00%) |  |
| **Sexual orientation** |  |  |  | 0.874 |
| Gay | 84 (74.34%) | 77 (72.64%) | 7 (100.00%) |  |
| Heterosexual | 8 (7.08%) | 8 (7.55%) | 0 (0.00%) |  |
| Bisexual | 12 (10.62%) | 12 (11.32%) | 0 (0.00%) |  |
| Other | 3 (2.65%) | 3 (2.83%) | 0 (0.00%) |  |
| DK/DA | 6 (5.31%) | 6 (5.66%) | 0 (0.00%) |  |
| **Country of birth:** |  |  |  | 0.759 |
| Spain | 37 (32.74%) | 34 (32.08%) | 3 (42.86%) |  |
| Other | 72 (63.72%) | 68 (64.15%) | 4 (57.14%) |  |
| DK/DA | 4 (3.54%) | 4 (3.77%) | 0 (0.00%) |  |
| **Level of studies:** |  |  |  | 0.670 |
| No studies |  |  |  |  |
| Complete primary school | 3 (2.65%) | 3 (2.83%) | 0 (0.00%) |  |
| Complete secondary school | 16 (14.16%) | 16 (15.09%) | 0 (0.00%) |  |
| Complete vocational studies | 33 (29.20%) | 30 (28.30%) | 3 (42.86%) |  |
| Complete Baccalaureate | 27 (23.89%) | 24 (22.64%) | 3 (42.86%) |  |
| Post-graduate studies (master, PhD, etc) | 30 (26.55%) | 29 (27.36%) | 1 (14.29%) |  |
| DK/DA | 4 (3.54%) | 4 (3.77%) | 0 (0.00%) |  |
| **Number of cohabitants:** | 2.00 [1.00;3.00] | 2.00 [1.00;3.00] | 3.00 [2.00;3.00] | 0.227 |
| **HIV positive** |  |  |  | 0.165 |
| Yes | 44 (38.94%) | 41 (38.68%) | 3 (42.86%) |  |
| No | 61 (53.98%) | 58 (54.72%) | 3 (42.86%) |  |
| I don't know my HIV status | 1 (0.88%) | 0 (0.00%) | 1 (14.29%) |  |
| I do not want to answer | 1 (0.88%) | 1 (0.94%) | 0 (0.00%) |  |
| DK/DA | 6 (5.31%) | 6 (5.66%) | 0 (0.00%) |  |
| **Take PrEP regularly** |  |  |  | 0.304 |
| Yes | 41 (59.42%) | 38 (58.46%) | 3 (75.00%) |  |
| No | 20 (28.99%) | 20 (30.77%) | 0 (0.00%) |  |
| DK/DA | 8 (11.59%) | 7 (10.77%) | 1 (25.00%) |  |
| **STI history in the last 12 months** |  |  |  |  |
| None | 52 (46.02%) | 49 (46.23%) | 3 (42.86%) | 1.000 |
| Syphilis | 20 (17.70%) | 20 (18.87%) | 0 (0.00%) | 0.585 |
| Chlamydia | 20 (17.70%) | 19 (17.92%) | 1 (14.29%) | 1.000 |
| Gonorrhea | 24 (21.24%) | 22 (20.75%) | 2 (28.57%) | 0.782 |
| Lymphogranuloma venereum | 1 (0.88%) | 1 (0.94%) | 0 (0.00%) | 1.000 |
| Genital herpes | 3 (2.65%) | 3 (2.83%) | 0 (0.00%) | 1.000 |
| Human papillomavirus | 5 (4.42%) | 5 (4.72%) | 0 (0.00%) | 1.000 |
| *Mycoplasma genitalium* | 7 (6.19%) | 7 (6.60%) | 0 (0.00%) | 1.000 |
| Hepatitis A | 0 (0.00%) | 0 (0.00%) | 0 (0.00%) | . |
| Hepatitis B | 0 (0.00%) | 0 (0.00%) | 0 (0.00%) | . |
| Hepatitis C | 3 (2.65%) | 2 (1.89%) | 1 (14.29%) | 0.228 |
| Other STIs | 6 (5.31%) | 6 (5.66%) | 0 (0.00%) | 1.000 |
| DK/DA | 3 (2.65%) | 1 (0.94%) | 2 (28.57%) | 0.014 |
| **Contact with a confirmed case of monkeypox:** |  |  |  | 0.570 |
| Yes | 28 (24.78%) | 25 (23.58%) | 3 (42.86%) |  |
| No | 63 (55.75%) | 60 (56.60%) | 3 (42.86%) |  |
| DK/DA | 22 (19.47%) | 21 (19.81%) | 1 (14.29%) |  |
| **Kind of contact:** |  |  |  |  |
| Person you take care of | 1 (3.57%) | 1 (4.00%) | 0 (0.00%) | 1.000 |
| Sexual contact | 11 (39.29%) | 9 (36.00%) | 2 (66.67%) | 0.543 |
| Shared food, utensils, or dishes | 11 (39.29%) | 9 (36.00%) | 2 (66.67%) | 0.543 |
| Shared clothes | 4 (14.29%) | 3 (12.00%) | 1 (33.33%) | 0.382 |
| Shared towels or bedding | 10 (35.71%) | 9 (36.00%) | 1 (33.33%) | 1.000 |
| Traveled together | 5 (17.86%) | 5 (20.00%) | 0 (0.00%) | 1.000 |
| Shared bathrooms | 9 (32.14%) | 8 (32.00%) | 1 (33.33%) | 1.000 |
| Physical contact | 22 (78.57%) | 20 (80.00%) | 2 (66.67%) | 0.530 |
| Other | 3 (10.71%) | 3 (12.00%) | 0 (0.00%) | 1.000 |
| DK/DA | 0 (0.00%) | 0 (0.00%) | 0 (0.00%) | . |
| **Risk of exposure to monkeypox virus at work:** |  |  |  | 1.000 |
| Yes | 10 (8.85%) | 10 (9.43%) | 0 (0.00%) |  |
| No | 90 (79.65%) | 84 (79.25%) | 6 (85.71%) |  |
| DK/DA | 13 (11.50%) | 12 (11.32%) | 1 (14.29%) |  |
| **Contact with pets:** |  |  |  | 1.000 |
| Yes | 32 (28.32%) | 30 (28.30%) | 2 (28.57%) |  |
| No | 78 (69.03%) | 73 (68.87%) | 5 (71.43%) |  |
| DK/DA | 3 (2.65%) | 3 (2.83%) | 0 (0.00%) |  |
| **Contact with exotic animals:** |  |  |  | 1.000 |
| Yes | 2 (1.77%) | 2 (1.89%) | 0 (0.00%) |  |
| No | 104 (92.04%) | 97 (91.51%) | 7 (100.00%) |  |
| DK/DA | 7 (6.19%) | 7 (6.60%) | 0 (0.00%) |  |
| **Traveled in the last 30 days:** |  |  |  | 1.000 |
| Yes | 53 (46.90%) | 50 (47.17%) | 3 (42.86%) |  |
| No | 57 (50.44%) | 53 (50.00%) | 4 (57.14%) |  |
| DK/DA | 3 (2.65%) | 3 (2.83%) | 0 (0.00%) |  |
| **Having received the Monkeypox vaccine** |  |  |  | 0.547 |
| Yes, vaccinated in childhood | 7 (6.19%) | 6 (5.66%) | 1 (14.29%) |  |
| Yes, vaccinated in the last 12 months | 13 (11.50%) | 12 (11.32%) | 1 (14.29%) |  |
| No | 75 (66.37%) | 71 (66.98%) | 4 (57.14%) |  |
| DK/DA | 18 (15.93%) | 17 (16.04%) | 1 (14.29%) |  |
| **Monkeypox Concern Level** |  |  |  | 0.833 |
| Extremely concerned | 40 (35.40%) | 38 (35.85%) | 2 (28.57%) |  |
| Moderately concerned | 40 (35.40%) | 37 (34.91%) | 3 (42.86%) |  |
| Somewhat concerned | 17 (15.04%) | 15 (14.15%) | 2 (28.57%) |  |
| Slightly concerned | 6 (5.31%) | 6 (5.66%) | 0 (0.00%) |  |
| Not at all concerned | 1 (0.88%) | 1 (0.94%) | 0 (0.00%) |  |
| DK/DA | 9 (7.96%) | 9 (8.49%) | 0 (0.00%) |  |
| **Self-perceived probability of getting monkeypox** |  |  |  | 0.778 |
| Very likely | 18 (15.93%) | 17 (16.04%) | 1 (14.29%) |  |
| Likely | 35 (30.97%) | 31 (29.25%) | 4 (57.14%) |  |
| Neither very nor unlikely | 29 (25.66%) | 27 (25.47%) | 2 (28.57%) |  |
| Unlikely | 13 (11.50%) | 13 (12.26%) | 0 (0.00%) |  |
| Very unlikely | 4 (3.54%) | 4 (3.77%) | 0 (0.00%) |  |
| DK/DA | 14 (12.39%) | 14 (13.21%) | 0 (0.00%) |  |

IQR, interquartile range; DK/DA, don't know/don't answer.

Behavioural characteristics of participants are shown in **Table 2**. The median number of sexual partners of participants over the previous 30 days was 5.00 (IQR: 1.00-10.00), of the total participants 42 (39.25%) had not used condoms during sexual intercourse over the previous month and 38 (33.63%) had had sex in exchange for money, gifts or favours. Furthermore, 29 (30.85%) had practiced chemsex in the previous 30 days and 3 (5.45%) had practiced slamming in the last month. Significant differences between participants with a positive or negative result for MPXV infection were found for the following variables: having practiced double penetration (vagina and anus) (P=0.004), slamming (p= 0.041), having met their sexual partners in music festivals in the last month (p=0.027) and taking their shirt off while partying (p= 0.033) (**Table 2**).

**Table 2.**
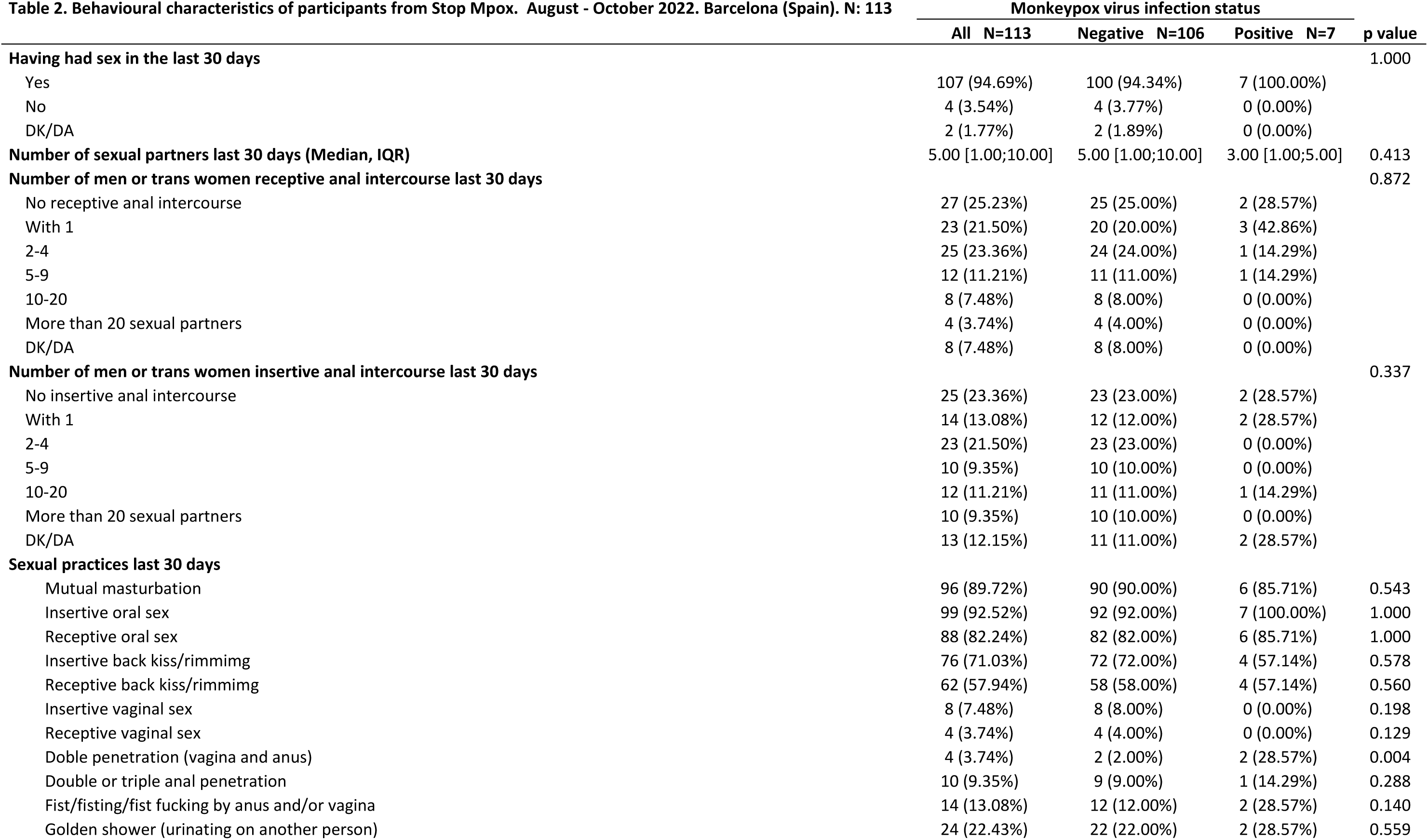
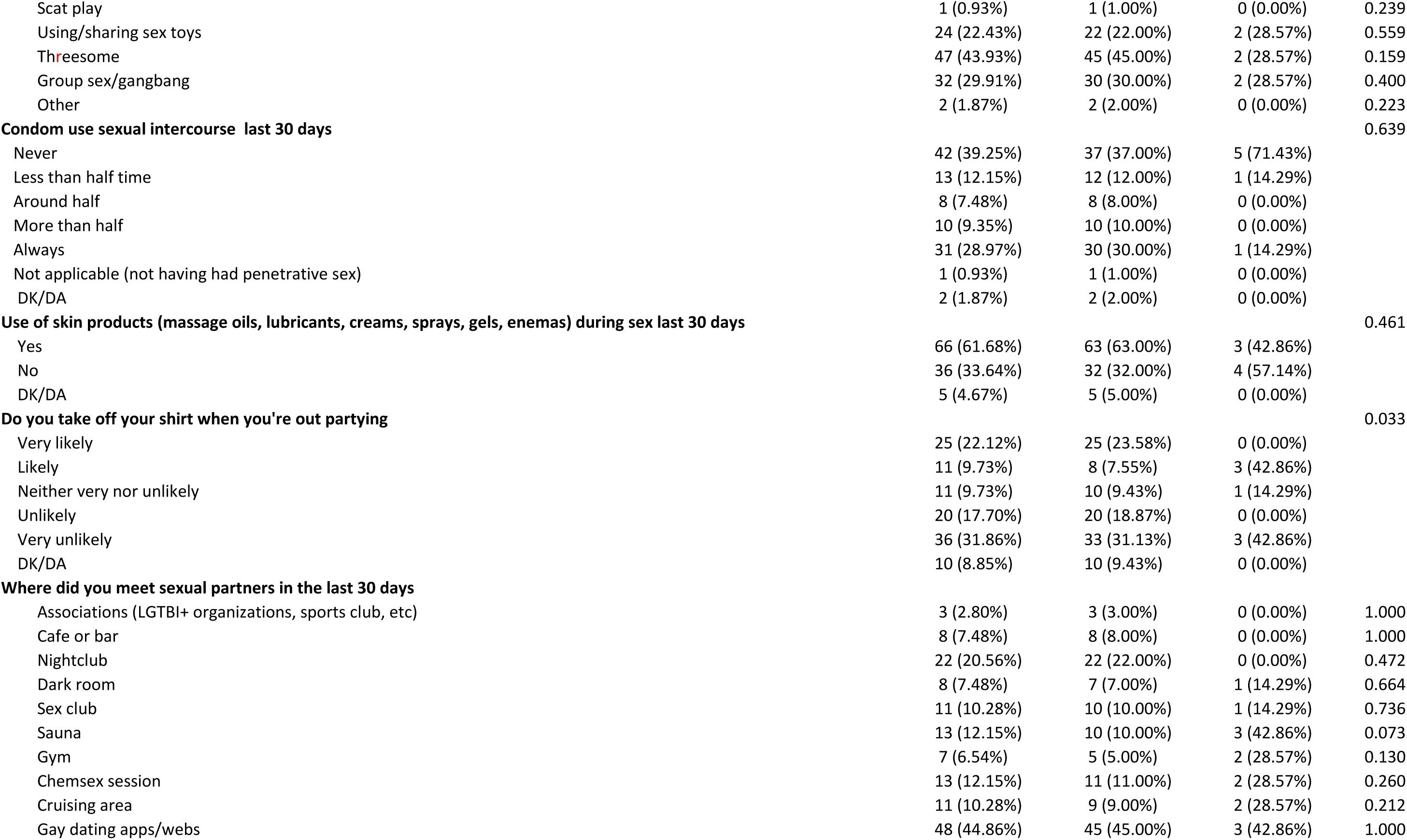
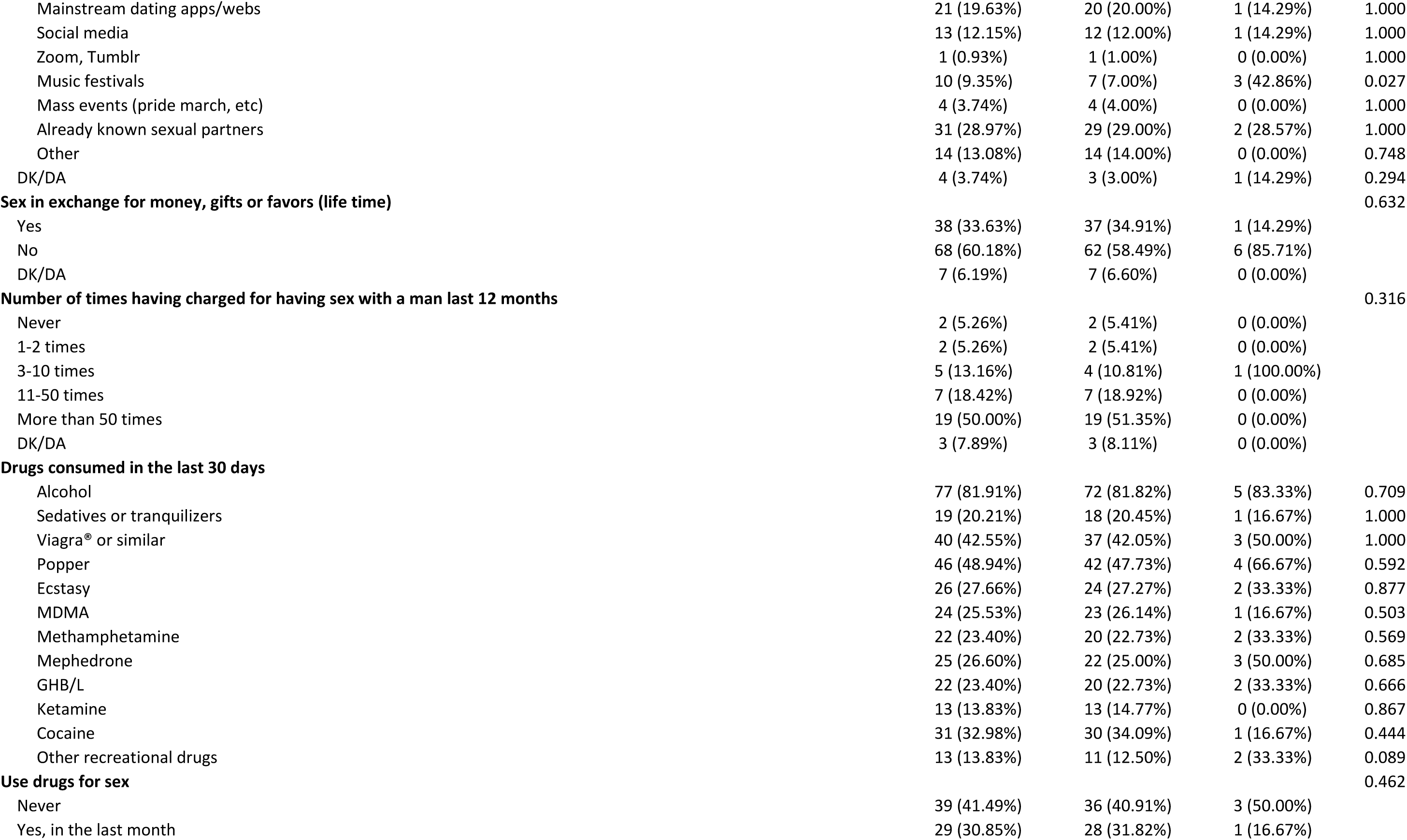
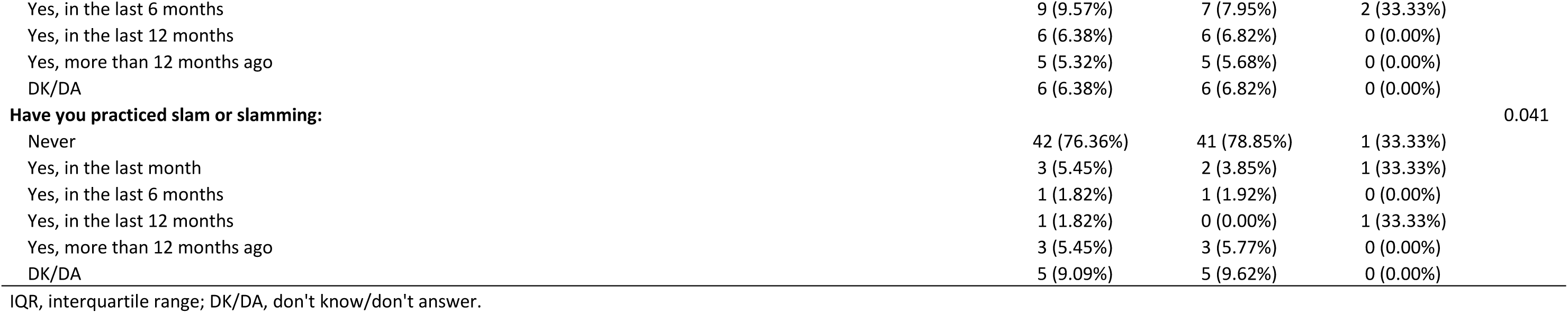
Behavioural characteristics of participants from Stop Mpox. August - October 2022. Barcelona (Spain). N: 113.

### MPXV prevalence

Analyses of 113 pharyngeal and 112 anal swabs, respectively, were performed in the reference laboratory. Eight positive MPXV results for seven individuals were detected and we estimated a total prevalence of 6.19% (95% CI: 1.75%-10.64%). All positive participants were cis gay men and prevalence in this group was 7.87% (95% CI: 2.27%-13.46%).

### Characteristics of individuals testing positive for MPXV

The characteristics of the participants with a positive MPXV result are shown in **Table 3**. Five participants tested positive in pharyngeal swabs, one in the anal swab and one in the pharyngeal and the anal swabs. PCR cycle thresholds (Ct) ranged from 24.85 to 38.06; and the viral load from 2,674 to 8,532,000 copies/mL.

**Table 3.** Results of MPXV infection among participants Stop MPX. August - October 2022. Barcelona (Spain). Samples were inoculated into cell cultures, which were assayed for viability over time (only viable samples with no contamination were followed up), the day of harvesting, the detection of cytopathic effect (CPE), the percentage of infected cells detected by flow cytometry (FACS) and the detection of immunofluorescence against vaccinia antigens (IF). Sample with asterisk denotes that it had to be sub-cultured again as primary culture exceeded 95 % of CPE which precluded proper FACS analysis.

| PCR Results |  |  |  |  | Cell Culture |  |  |  |  |  |  | Symptoms |
| --- | --- | --- | --- | --- | --- | --- | --- | --- | --- | --- | --- | --- |
| Individual | Location | Result | Ct | CV<br>(Copies/mL) | Culture<br>viability | Day<br>of Harvesting | CPE | IF | FACS | Ct | TCID<sub>50</sub>/ml |  |
| **2** | Pharyngeal | Positive | 34.9 | 18300 | No | 4 | N/A | N/A | N/A | N/A | N/A | No reported symptoms |
|  | Anal | Negative | - | - | N/A | N/A | N/A | N/A | N/A | N/A | N/A |  |
| **40** | Pharyngeal | Positive | 30.1 | 347000 | Yes | 8 | Positive | Positive | 8.41% | 21.96 | 10<sup>4,3</sup> | No symptoms before testing.<br>After testing the participant reported:<br>Fever, exhaustion, sore throat and a skin lesion |
|  | Anal | Negative | - | - | N/A | N/A | N/A | N/A | N/A | N/A | N/A |  |
| **64** | Pharyngeal | Positive | 37.09 | 4827 | Yes | 8 | Negative | Negative | Negative | Negative | Negative | No available information |
|  | Anal | Negative | - | - | N/A | N/A | N/A | N/A | N/A | N/A | N/A |  |
| **66** | Pharyngeal | Positive | 24.85 | 8532000 | Yes | 7 | Positive | Positive | 74.2%* | 20.42 | 10<sup>4,8</sup> | Before testing: A swollen inguinal lymph node |
|  | Anal | Positive | 35.35 | 13960 | No | 2 | N/A | N/A | N/A | N/A | N/A |  |
| **72** | Pharyngeal | Negative | - | - | N/A | N/A | N/A | N/A | N/A | N/A | N/A | Before testing: Fever, exhaustion and<br>a skin lesion in the genital area |
|  | Anal | Positive | 38.06 | 2674 | Yes | 7 | Positive | Positive | 3.75% | 35.98 | <10<sup>1</sup> |  |
| **81** | Pharyngeal | Positive | 36.99 | 5126 | Yes | 8 | Negative | Negative | Negative | Negative | Negative | No reported symptoms |
|  | Anal | Negative | - | - | N/A | N/A | N/A | N/A | N/A | N/A | N/A |  |
| **95** | Pharyngeal | Positive | 36.79 | 5825 | Yes | 14 | Positive | Positive | 3.18% | 25.88 | 10<sup>2,8</sup> | Skin lesions in the genital area |
|  | Anal | Negative | - | - | N/A | N/A | N/A | N/A | N/A | N/A | N/A |  |
| **112** | Pharyngeal | Negative | - | - | Yes | 8 | Negative | Negative | Negative | Negative | Negative | - |
| **113** | Pharyngeal | Negative | - | - | Yes | 8 | Negative | Negative | Negative | Negative | Negative | - |
| **Control +** | Viral stock | N/A | - | - | Yes | 7 | Positive | Positive | Positive | 19.21 | 10<sup>6,8</sup> | - |
| **Control -** | Culture media | N/A | - | - | Yes | 8 | Negative | Negative | Negative | Negative | Negative | - |

Ct: Cycle threshold. CPE: Cytopathic Effect. IF: Immuno fluorescence. FACS: Fluorescence Assisted Cell Sorting. Samples assayed to isolate infectious viruses and results obtained.

We aimed to isolate viable virus from the pharyngeal and anal swab samples with reported PCR positive results for MPXV. We included two undetectable samples along with media as negative controls. As a positive control, we used a viral stock previously isolated during the 2022 summer belonging to the same outbreak and geographical location. We cultured the samples and followed them up for 14 days or until cytopathic effect (CPE) was detected under the microscope when infected cells were assessed for the detection of viral antigens using an anti-vaccinia polyclonal antibody (pAb) by fluorescence-activated cell sorting (FACS) and confocal microscopy. Although two viral cultures had to be discarded due to the presence of contaminant microorganisms, we were able to follow up ten for 14 days (**Table 3**). Three out of the six PCR positive samples successfully cultured over time (40, 66 and 95) were unambiguously positive, not only for CPE at the indicated day of harvesting, but also for staining for specific anti-vaccinia pAb detected by FACS and confocal microscopy. Sample 72 was at the limit of positivity (TCID_50_<10) and was not considered as a positive (**Table 3** and **Supplemental Figure 1**). The Ct values of samples 40, 66 and 95 ranged from 24.85 to 36.79, indicating that samples with low viral load, such as the one collected by individual 95, were competent for cellular infection *in vitro* (**Table 3**). We also tested the recovered viral stocks by PCR, which yielded positive results for MPXV, and we sequenced them along with the positive control viral stock, which confirmed the specificity of the MPXV presence in cell cultures, while we detected no signal in the negative control. Furthermore, infectivity was measured as tissue culture infectious doses per ml (TCID_50_/mL; **Table 3**). Negative samples and media were negative for all of the analyses, while the inoculation with a viral stock resulted in positive CPE and viral antigen detection by FACS and confocal microscopy (**Table 3** and **Supplemental Figure 1**). These results demonstrated a clear propagation of infectious MPXV from at least three out of the six (3/6) samples that were successfully cultured over time from the original 8 MPXV positive samples identified.

Eight anal samples yielded an inconclusive PCR result, as no human DNA (myostatine gene) was detected due to the presence of inhibitors or because the sample was not correctly collected.

Regarding presentation of symptoms, two (2/6) positive-testing participants reported having no symptoms before testing, or 21 days after. One (1/6) had no symptoms before testing and reported having fever, exhaustion, sore throat and a skin lesion in the 21 days following testing positive. Three (3/6) participants reported the following symptoms before testing: a swollen inguinal lymph node, fever, exhaustion and a skin lesion but none of them connected these symptoms with MPXV infection. There was no information available regarding one of the participants with a positive MPXV result (**Table 3**). It is of note that viable MPXV viruses were obtained from individuals reporting symptoms, although one had no symptoms before testing. These results highlight the fact that pre-symptomatic phases of infection have the potential to promote ongoing viral transmission events in the community.

### Acceptability and feasibility of the self-sampling intervention

In relation to the acceptability and usability of the self-sampling procedure, 88 (77.87%) and 90 (79.64%) participants considered that the self-sampling procedure was easy or very easy for pharyngeal and anal swabs respectively; and 99 (87.61%) and 98 (86.72%) agreed or agreed very strongly with the statement “I reckon I have collected the pharyngeal sample correctly” and “I reckon I have collected the anal sample correctly”, respectively (**Table 4**). In addition, 98 (86.73%) participants were satisfied or very satisfied with the self-sampling screening intervention and 103 (99.0%) agreed or agreed very strongly that they would recommend participating in the intervention to a friend. The most commonly identified advantages of the intervention by participants were: (i) privacy and confidentiality (75.22%) and (ii) that the test was free (74.34%) (**Table 4**). The most preferred place to repeat the MPXV test if necessary was a community-based centre (53.10%) and most preferred performing self-sampling at home (48.67%) compared to attending a health care setting (41.59%). Significant differences were found between participants with a positive and negative result for MPXV; all participants testing positive chose to repeat the test at a community-based centre compared to participants with a negative result (50.00%) (p value: 0.019) (**Table 4**). These results do not only emphasize the acceptance of self-collected samples, but also the feasibility of using this strategy for downstream laboratory analyses involving viral isolation techniques.

**Table 4.** Usability and acceptability of the self-sampling intervention to detect MPVX. Stop Mpox. August - October 2022. Barcelona (Spain). N: 113

|  | Monkeypox virus infection status |  |  | p value |
| --- | --- | --- | --- | --- |
|  | All N=113 | Negative N=106 | Positive N=7 |  |
| **Difficulty level pharyngeal self-sampling** |  |  |  | 0.950 |
| Very easy | 57 (50.44%) | 53 (50.00%) | 4 (57.14%) |  |
| Easy | 31 (27.43%) | 28 (26.42%) | 3 (42.86%) |  |
| Neither easy nor difficult | 8 (7.08%) | 8 (7.55%) | 0 (0.00%) |  |
| Difficult | 7 (6.19%) | 7 (6.60%) | 0 (0.00%) |  |
| Very difficult | 1 (0.88%) | 1 (0.94%) | 0 (0.00%) |  |
| DK/DA | 9 (7.96%) | 9 (8.49%) | 0 (0.00%) |  |
| **Difficulty level anal self-sampling** |  |  |  | 0.937 |
| Very easy | 61 (53.98%) | 57 (53.77%) | 4 (57.14%) |  |
| Easy | 29 (25.66%) | 27 (25.47%) | 2 (28.57%) |  |
| Neither easy or difficult | 9 (7.96%) | 9 (8.49%) | 0 (0.00%) |  |
| Difficult | 5 (4.42%) | 5 (4.72%) | 0 (0.00%) |  |
| Very difficult | 0 (0.00%) | 0 (0.00%) | 0 (0.00%) |  |
| DK/DA | 9 (7.96%) | 8 (7.55%) | 1 (14.29%) |  |
| **I trust that I have collected the pharyngeal sample correctly** |  |  |  | 0.685 |
| Agree strongly | 72 (63.72%) | 68 (64.15%) | 4 (57.14%) |  |
| Agree | 27 (23.89%) | 24 (22.64%) | 3 (42.86%) |  |
| Neither very nor disagree | 5 (4.42%) | 5 (4.72%) | 0 (0.00%) |  |
| Disagree | 0 (0.00%) | 0 (0.00%) | 0 (0.00%) |  |
| Disagree strongly | 1 (0.88%) | 1 (0.94%) | 0 (0.00%) |  |
| DK/DA | 8 (7.08%) | 8 (7.55%) | 0 (0.00%) |  |
| **I trust that I have collected the anal sample correctly** |  |  |  | 0.610 |
| Agree strongly | 73 (64.60%) | 69 (65.09%) | 4 (57.14%) |  |
| Agree | 25 (22.12%) | 22 (20.75%) | 3 (42.86%) |  |
| Neither very nor disagree | 5 (4.42%) | 5 (4.72%) | 0 (0.00%) |  |
| Disagree | 1 (0.88%) | 1 (0.94%) | 0 (0.00%) |  |
| Disagree strongly | 0 (0.00%) | 0 (0.00%) | 0 (0.00%) |  |
| DK/DA | 9 (7.96%) | 9 (8.49%) | 0 (0.00%) |  |
| **Level of satisfaction self-sampling intervention to detect monkeypox** |  |  |  | 1.000 |
| Very satisfied | 66 (58.41%) | 61 (57.55%) | 5 (71.43%) |  |
| Satisfied | 32 (28.32%) | 30 (28.30%) | 2 (28.57%) |  |
| Neither satisfied nor unsatisfied | 6 (5.31%) | 6 (5.66%) | 0 (0.00%) |  |
| Unsatisfied | 0 (0.00%) | 0 (0.00%) | 0 (0.00%) |  |
| Very unsatisfied | 0 (0.00%) | 0 (0.00%) | 0 (0.00%) |  |
| DK/DA | 9 (7.96%) | 9 (8.49%) | 0 (0.00%) |  |
| **Would you repeat self-sampling to detect monkeypox** |  |  |  | 0.612 |
| Agree strongly | 84 (74.34%) | 79 (74.53%) | 5 (71.43%) |  |
| Agree | 16 (14.16%) | 14 (13.21%) | 2 (28.57%) |  |
| Neither very nor disagree | 4 (3.54%) | 4 (3.77%) | 0 (0.00%) |  |
| Disagree | 0 (0.00%) | 0 (0.00%) | 0 (0.00%) |  |
| Disagree strongly | 1 (0.88%) | 1 (0.94%) | 0 (0.00%) |  |
| DK/DA | 8 (7.08%) | 8 (7.55%) | 0 (0.00%) |  |
| **Would you recommend self-sampling to detect monkeypox to a friend** |  |  |  | 0.087 |
| Agree strongly | 91 (80.53%) | 87 (82.08%) | 4 (57.14%) |  |
| Agree | 12 (10.62%) | 9 (8.49%) | 3 (42.86%) |  |
| Neither very nor disagree | 0 (0.00%) | 0 (0.00%) | 0 (0.00%) |  |
| Disagree | 0 (0.00%) | 0 (0.00%) | 0 (0.00%) |  |
| Very disagree | 1 (0.88%) | 1 (0.94%) | 0 (0.00%) |  |
| DK/DA | 9 (7.96%) | 9 (8.49%) | 0 (0.00%) |  |
| **Consider self-sampling to detect monkeypox as a good intervention for monkeypox screening** |  |  |  | 0.089 |
| Agree strongly | 86 (76.11%) | 82 (77.36%) | 4 (57.14%) |  |
| Agree | 16 (14.16%) | 14 (13.21%) | 2 (28.57%) |  |
| Neither very nor disagree | 3 (2.65%) | 2 (1.89%) | 1 (14.29%) |  |
| Disagree | 0 (0.00%) | 0 (0.00%) | 0 (0.00%) |  |
| Disagree strongly | 0 (0.00%) | 0 (0.00%) | 0 (0.00%) |  |
| DK/DA | 8 (7.08%) | 8 (7.55%) | 0 (0.00%) |  |
| **Self-perceived advantages of self-sampling to detect monkeypox** |  |  |  |  |
| Privacy and confidentiality | 85 (75.22%) | 78 (73.58%) | 7 (100.00%) | 0.525 |
| More convenience since you don't have to go to the medical center | 80 (70.80%) | 73 (68.87%) | 7 (100.00%) | 0.326 |
| The test is free | 84 (74.34%) | 77 (72.64%) | 7 (100.00%) | 0.527 |
| No prescription needed | 61 (53.98%) | 57 (53.77%) | 4 (57.14%) | 1.000 |
| No need to give explanations | 76 (67.26%) | 71 (66.98%) | 5 (71.43%) | 1.000 |
| It contributes to normalize the monkeypox test | 61 (53.98%) | 57 (53.77%) | 4 (57.14%) | 1.000 |
| Allows me to take control of my health regarding monkeypox | 73 (64.60%) | 69 (65.09%) | 4 (57.14%) | 0.769 |
| DK/DA | 2 (1.77%) | 2 (1.89%) | 0 (0.00%) | 1.000 |
| **Self-perceived disadvantages of self-sampling to detect monkeypox** |  |  |  |  |
| That the test requires the introduction of a swab orally | 14 (12.39%) | 13 (12.26%) | 1 (14.29%) | 1.000 |
| That the test requires the introduction of a swab via the anus | 14 (12.39%) | 13 (12.26%) | 1 (14.29%) | 1.000 |
| Not having the result at the moment | 49 (43.36%) | 45 (42.45%) | 4 (57.14%) | 0.876 |
| Not having emotional support when receiving the result | 10 (8.85%) | 9 (8.49%) | 1 (14.29%) | 0.810 |
| The time to receive the result is too long | 16 (14.16%) | 13 (12.26%) | 3 (42.86%) | 0.091 |
| Other | 5 (4.42%) | 5 (4.72%) | 0 (0.00%) | 1.000 |
| DK/DA | 22 (19.47%) | 20 (18.87%) | 2 (28.57%) | 0.855 |
| **Preferred location for repeat testing if necessary** |  |  |  |  |
| Health care centre | 47 (41.59%) | 42 (39.62%) | 5 (71.43%) | 0.238 |
| Community based centre | 60 (53.10%) | 53 (50.00%) | 7 (100.00%) | 0.019 |
| Self-sampling at home | 55 (48.67%) | 51 (48.11%) | 4 (57.14%) | 1.000 |
| DK/DA | 8 (7.08%) | 8 (7.55%) | 0 (0.00%) | 1.000 |
| DK/DA, don't know/don't answer. |  |  |  |  |

## Discussion

Our study provides evidence that there are Mpox cases that remain undiagnosed because patients have no symptoms, or because they have mild unrecognized Mpox symptoms^8, 9^. The estimated prevalence of this viral infection was 6.19% for all individuals and 7.87% for cis gay men. Three out of the 7 participants who tested positive for MPXV through our study did not have any symptom before testing, while three of them had symptoms that were easily confused with STIs that cause skin rashes or mucosal lesions, such as genital herpes, syphilis, acuminate condyloma and chancroid among others; or even COVID-19 (which causes fever and exhaustion). Our results are highly relevant for transmission dynamics as we also show that these particular cases had viable infectious viruses in cell culture. We were able to identify replication-competent virus particles in three out of six (3/6) MPXV positive individuals. This indicates that transmission from these cases is possible. One of them did not present symptoms before testing and the other two did not recognize their symptoms as indicative of MPXV infection. As positive MPXV cases were not aware of their infection, without our self-sampling intervention they would not have attended a health care setting, or get diagnosed, and consequently they would not have self-isolated and we would not have carried out contact tracing. Therefore, if they had not known their infection status and as they presented no symptoms, these individuals would have continued to spread the infection unknowingly.

Tarin et al^6^ proposed skin-to-skin contact rather than the respiratory route as the dominant mode of MPXV transmission outside countries where the virus is endemic based on the history of sexual exposure, predominant anogenital skin lesions, and higher viral loads in skin than throat swabs. Here, we confirm that pharyngeal swabs allow for the isolation of viable viruses. As, MPXV has been isolated from semen^18–20^ these findings further corroborates the role of sexual transmission of MPXV during the 2022 outbreak.

All positive cases in our study were found in cis gay men. The global spread of MPXV among GBMSM has shown that sexual contact is a new mode of MPXV transmission and it has implications for infection control, contact tracing policies, education and prevention strategies, and clinical management of individuals accessing sexual health services^18, 21^.

Our results suggest that restricting testing only to individuals reporting symptoms compatible with MPXV infection may not be enough to contain an ongoing outbreak. In areas with high community transmission, screening for MPXV in pharyngeal and anal swabs should be offered to those GBMSM at risk of acquiring STIs. The establishment of health policies to promote early diagnosis and linkage to care as well as contact tracing is crucial to contain the current world-wide outbreak.

Although Mpox cases have been reported among TW and non-binary individuals^22^, no cases among TW have been detected in our study or previous studies performed in Spain^4, 23^. Nevertheless, TW are highly vulnerable to HIV and other STI infections; and in the case of HIV the WHO has recognized the high vulnerability and specific health needs of transgender people with the consequent need for a distinct and independent status in the global HIV response^24^. The disparities in MPXV prevalence between TW and GBMSM could be explained by distinct sexual networks across populations without shared transmission, which is similar to HIV transmission patterns described among TW, their sexual partners and GBMSM^25^.

This study gained access to GBMSM and TW at high risk of HIV and STIs and the results show the benefits of working together with community organizations. This collaboration needs to take place in the conceptualization of the study, elaboration of the messages, dissemination of the intervention and when facilitating access to the target population.

Ubals *et al.* have recently described that MPXV diagnostic tests with both self-collected swabs and physician-collected swabs have shown a similarly high accuracy and yield similar Ct values in both samples^17^. We demonstrated that a self-sampling intervention for MPXV screening in collaboration with a community centre is feasible and acceptable. It resulted in high levels of satisfaction and willingness to participate from the target population and most of participants considered it easy or very easy to self-collect the samples. Moreover, it also allowed us to perform highly sensitive laboratory techniques downstream, such as viral isolation in cell culture.

Linkage to care is often challenging in self-sampling strategies. We obtained high percentages of confirmation and linkage to care since 6 out of 7 (85.7%) MPXV positive participants self-reported having been linked to care. The linkage to care rate obtained is similar to previous studies on self-sampling strategies for HIV screening also addressed to GBMSM^14, 15^ and comparable to the percentage of individuals with a reactive screening test for HIV who were linked to care in a network of community-based services, which offer voluntary counselling and testing for HIV in Spain^26^. Follow-up of participants with a positive result should be reinforced to improve rates of linkage to care.

Our study presents several limitations. Firstly, the study population is not representative of GBMSM and TW in Catalonia as we used an opportunistic sample. Secondly, while self-collected anal swabs have been described as a feasible, valid, and acceptable alternative for men who have sex with men and women attending STI clinics^27^, an inconclusive PCR result was obtained in 7% in these samples compared to 0% in pharyngeal swabs. Thirdly, we analysed both pharyngeal and anal samples, but for logistical reasons we did not include seminal samples, which, although they may not affect the overall prevalence of MPXV, may be necessary to better define the potential routes of transmission. Finally, the small number of TW tested within this study could have precluded the detection of positive cases in this group.

In conclusion, our findings have important public health implications, particularly for MPXV infection prevention and control policies. We have shown that MPXV infection is present among asymptomatic individuals and among vulnerable populations. We also demonstrate that MPXV symptoms can overlap and be confused with other diseases, such as other STIs. Moreover, we showed that asymptomatic or very mild symptomatic patients might be able to transmit the infection to others, as we were able to isolate replication-competent viruses from pharyngeal and anal swabs, according to previous studies^9^. First of all, educational interventions are needed to familiarize the members of vulnerable populations with the nature of MPXV symptoms and eradicate the associated stigma in order to increase awareness and health care seeking behaviour in these populations. Secondly, in an epidemic scenario, early diagnosis by means of screening strategies should be aimed not only at suspected clinical cases and direct contacts, but also at all GBMSM at high risk of contracting Mpox, regardless of their symptoms. Community-based self-sampling tools can be an acceptable and effective to increase early diagnosis and the eventual isolation of infectious cases. On the other hand, heath care workers in STI clinics, primary care, and emergency rooms in other health care settings should be aware of the variety of Mpox symptoms and the possibility of asymptomatic cases before excluding Mpox as a potential diagnosis. Finally, stigma and discrimination in the most affected group, GBMSM, should be addressed to warrant equitable access to diagnosis, treatments and vaccines. More data are needed to better establish the attributable risk of asymptomatic infections in the transmission of MPXV in an outbreak, including seminal transmission.

## Online Methods

### Study design and setting

We implemented a transversal non-randomized study offering free self-sampling kits for MPXV testing through a collaborating community centre that offers voluntary counselling and testing for HIV (STOP, Barcelona, Spain). The field coordinator communicated test results to participants by a phone call.

### Study population and recruitment

The study targeted two different key populations: GBMSM and TW, over 18 years old, with no symptoms of MPXV infection and considered at high risk of contracting Mpox. High risk was defined as: GBMSM and TW who are sex workers and/or chemsex users and/or who practice group sex and/or are HIV positive or are PrEP users. The study was disseminated through Instagram, Facebook, Whatsapp and the community centre website via intermittent campaigns. Participants with eligible criteria were invited to attend the collaborating community centre to get tested for MPXV.

The community centre staff briefly explained the project to potential participants and obtained the signed informed consent on paper. Participants answered a self-completed paper survey on behaviour and self-collected the samples.

Data were collected prospectively from August to October 2022 in Barcelona, Spain.

### Data collection instrument

The following data was collected through the survey: Year and country of birth, sex at birth, gender identity, sexual orientation, level of studies, monthly salary, sex work, having used drugs in the last three months, chemsex in the last three months. Recent Sexual History and risky Practices (<30 days): Number of sexual partners, sexual practices, condom use. Use of PrEP. History of smallpox vaccination. Potential MPX exposures within the last 30 days: Close contact with a MPX infected case, contact with animals, history of travelling, occupational exposure. History of STI diagnosis. HIV status, PrEP use. Risk perception towards MPXV infection (likelihood of infection, level of concern). Acceptability of the pilot intervention: level of satisfaction, willingness to repeat the experience, likelihood of recommending it to a friend, perceived advantages and disadvantages and preferred way to repeat the MPXV test.

We used RedCAP (REDCap systems, Vanderbilt University, US) to collect data of participants and create an *ad hoc* online data base. We carried out the data entry of the survey data at the coordinating centre.

#### Self-sampling kits

The self-sampling kits included an anal and a pharyngeal swab (Standard Swab, Deltalab, Rubí, Spain), pre-labelled swab containers and a brochure with detailed instructions with pictures explaining how to get the samples. A video with the instructions of sample collection was available on YouTube and was accessible through a QR code included in the brochure. Participants were able to contact the field coordinator by phone or email if they have any doubt. After obtaining the sample, the swabs were immediately placed in 1 ml of transport medium (Standard Swab, Deltalab, Rubí, Spain) and samples were stored at 4 °C. A parcel courier service provided the secondary and tertiary containers, and samples were transported at 4°C to the reference laboratory (Microbiology Department, LCMN, Germans Trias i Pujol University Hospital).

#### Delivering test results and follow-up of participants with a positive result

The field coordinator delivered test results by a phone call. All participants with a positive result for MPXV infection were asked to attend their General Practitioner (GP) or a STI Clinic for confirmation and were advised on isolation measures. After three weeks, these participants were contacted by phone to enquire if they had had any symptom before testing them within the following 21 days.

#### Lab analysis

##### PCR assays

We analyzed all samples for the detection of MPXV DNA with a real-time PCR-based assay (qualitative and quantitative) at the reference laboratory. We performed nucleic acid extraction using the Seegene StarLet platform (Hamilton Company, Reno, US), according to manufacturer’s instructions. We carried out quantitative PCR (qPCR) using the LightMix Modular Monkeypox Virus assay (TIB MolBiol, Berlin, Germany) with LightMix Modular MSTN Extraction Control (TIB MolBiol, Berlin, Germany) as the internal control. We used a thermocycler QuantStudioTM 5 Real-Time PCR System (Applied Biosystems) to amplify an 89 bp-long fragment of the myostatin gene of vertebrates as an internal control and 106 bp-long fragment of the J2L/J2R gene from MPXV. We used Applied Biosystems Interpretive Software for detection and data analysis. To determine copy number per mL we used a linear dilution series of a quantified MPXV DNA standard (AMPLIRUN® Monkeypox virus DNA control, Vircell Spain SLU, Santa Fe, Granada, Spain). The calibration curve was composed of 5 points containing 1,000,000, 100,000, 10,000, 1,000 and 100 copies/mL (6.00, 5.00, 4.00, 3.00 and 2.00 Log_10_ copies/mL, respectively), and for each point we analysed in three replicates together with negative and positive controls. To calculate the MPXV DNA in study samples we extrapolated Ct data from the standard curve.

##### Cells, viral isolation and titration

We cultured Vero E6 cells (ATCC CRL-1586) at 37°C and 5% CO_2_ in Dulbecco’s modified Eagle medium (DMEM; Invitrogen) supplemented with 5% foetal bovine serum (Invitrogen), 100 U/mL penicillin and 100 µg/mL streptomycin (all ThermoFisher Scientific).

To create the positive control in this study MPXV stock was isolated in August 2022 from a skin lesion swab from a patient diagnosed with Mpox illness. Briefly, we cultured Vero E6 cells in T25 culture flasks (25 cm2) at 1.5×10^6^ cells and inoculated them with 1 mL of the liquid sample, for 1 h at 37°C and 5% CO_2_.Then we added, 4 ml of 2% FCS-supplemented DMEM containing 100 U/mL penicillin, 100 µg/mL streptomycin and 2,5 µg/mL amphotericin B (all from ThermoFisher Scientific). We maintained cells in incubation and assessed them daily for cytopathic effect (CPE) in order to be able to harvest the supernatant, which was centrifuged at 410 g for 5 min to remove cell debris and stored at −80°C. We propagated the virus for two passages and collected the supernatant. We titrated the viral stock and confirmed the infection by the presence of viral antigens using antibodies as described below.

##### Viral isolation from clinical samples

We inoculated pharyngeal or anal swab samples that had either a positive detection of MPVX DNA by PCR (n=8) or negative results (n=2) into T25 culture flasks with Vero E6 cells as we have described previously. As a positive control we employed the MPVX stock we had previously isolated, and as a negative control we included mock-treated cells. We assessed viral cultures daily and kept them for 14 days or until 50% CPE was observed. In cases where we detected CPE, we harvested the supernatants, centrifuged them at 410g for 5 min to remove cell debris and stored them at −80°C. We washed cells from these positive cultures once with PBS, detached them using 0.5% EDTA trypsin, collected and resuspended them in 0.5 mL of PFA 4% (Merck) for fixation. If we detected no CPE, the cells remained in culture until reaching a confluent state, when we passed half of the cells with supernatant from the previous culture to new flasks and added antibiotics and amphotericin B. We discarded any cultures with undesired microorganism growth and did not consider them for the analysis. We followed the cultures for up to 14 days.

##### Immunostaining and FACS analysis

We resuspended fixed cells from positive cultures in 100 µL of Permeabilization Medium (Invitrogen) with a rabbit polyclonal antibody vaccinia virus (Abcam, ab35219) at 1:2000 dilution (2 µg/mL) and incubated them for 20 min at room temperature in darkness. We removed the primary antibody by washing with blocking buffer (PBS, 5% FBS). Then we performed a secondary incubation with a goat anti-rabbit IgG H&L Alexa Fluor® 488 antibody (Abcam, ab150077), which we added at 1:1000 dilution (2 µg/mL) and cells were incubated for 20 min at room temperature in darkness. After a PBS wash, we resuspended cells with 300 µL of PBS 1% PFA. To analyze the samples we used a FACSCalibur (Becton-Dickinson) and CellQuest and FlowJo v10.6.1 software to evaluate collected data.

##### Confocal microscopy

We prepared microscopy slides (cytospins) with 100 µL of previously fixed and immunostained Vero E6 cells, using EZ Double Cytofunnel (Fisher Scientific) and mounted samples onto slides with Fluoromount-G™ Mounting Medium, with DAPI (Life Technologies). We used a confocal LSM710 microscope and a 63X objective at the IGTP Microscopy Facility to image the cells.

##### Viral DNA extraction

We carried out viral DNA extraction with the QiaAmp Viral RNA Mini kit (Qiagen), although this kit has been optimized for the extraction of RNA, it is possible to obtain DNA in parallel according to the manufacturer’s instructions. We extracted viral DNA from 140µL of cellular culture supernatants to perform MPXV PCR as previously described.

#### Titration of viral isolates from clinical samples

We titrated the viral supernatants collected in 1/10 dilutions using 96-well-plate containing 30.000 Vero E6 cells per well and inspected the plates at the microscope for CPE 6 days post-infection. We were able to calculate the TCID_50_ per mL by inferrence from the number of positive and negative wells using the Reed & Muench method^28^.

#### Whole genome sequencing

Starting from DNA extracted from culture supernatants, we amplified the whole MPXV genome by adapting the amplicon tiling approach described by Welkers *et al*.^29^ using Q5™ Hot Start High-Fidelity 2× Master Mix (New England Biolabs, USA) with the following cycling conditions: 30 s at 98°C, and then 35 cycles for 10 s at 98°C 10 sec and 5 min at 65°C. We prepared sequencing libraries using the Rapid Barcoding Kit 96 from Oxford Nanopore Technologies (ONT, UK), which was pooled and loaded onto a R9.4.1 flow cell and sequenced in for 72 hours on a MinION Mk1C. We processed a negative control along with the samples in order to monitor the whole process.

#### Bioinformatics analysis

We analysed raw sequencing data using a custom Nextflow pipeline. In summary we trimmed reads for length and quality using NanoFilt (v2.8.0)^30^, then aligned them with a MPXV reference genome (MT903344.1) using minimap2 (v2.24-r1122)^31, 32^ We built draft consensus sequences using bcftools (v1.15). Finally, we used Nextclade (v2.5.0)^33, 34^ to assess consensus quality and assign MPXV lineages.

#### Statistical analysis

MPXV infection prevalence was estimated by calculating the proportion of individuals with a positive result over the total of individuals with a returned and valid sample. Confidence interval of 95% was calculated.

We carried out descriptive analysis to compare socio-demographic characteristics, risk behaviour variables, and previous STI diagnoses between participants with a positive and negative MPVX test result. Continuous variables were expressed as medians and IQRs. Categorical variables were summarised as absolute values and proportions. Qualitative variables were compared using Pearson’s χ2 test. We made quantitative variable comparisons between two or more groups using non-parametric tests (Kruskal-Wallis) and no imputation was made for missing data. For all analysis, a significance level of 5% was considered. All analyses were done using R version 4.0.5.

#### Ethical considerations

All identifying data collected was encrypted. Confidentiality was guaranteed in accordance with the provisions of the Regulation (EU) 2016/679 of the European Parliament and of the Council of 27 April 2016 and the new national Organic Law of Protection of Personal Data (3/2018, 5 December, Data Protection and Digital Rights Act). We provided written information about the study to all participants and they had the opportunity to ask questions and clarify queries with the study coordinator by email or phone. The Ethical Committee of the Germans Trias i Pujol Hospital approved the study protocol (PI-22-195). The biological biosafety committee of the Germans Trias i Pujol Research Institute approved the execution of MPXV experiments at the BSL3 laboratory of the Centre for Comparative Medicine and Bioimage (CMCiB, protocol number CSB-22-011-M1).

## Data Availability

All data produced in the present study are available upon reasonable request to the authors

## Tables and figures

Supplementary Figure 1: Representative results from samples assayed to isolate infectious MPVX. A. Optical microscopy images of Vero E6 cultures inoculated with swab samples. Images were taken at the day post infection (dpi) indicated in the top left part of the images. Scale bars correspond to 100 µm. B. Confocal microscopy images of cells where swab samples were grown after intracellular staining with the anti-vaccinia pAb revealed with an Alexa 488 secondary antibody from the cultures. DAPI staining is shown in blue (nuclei) and α-Vaccinia is shown in green. Scale bars correspond to 20 µm. C. Percentage of positive cells detected by FACS after intracellular staining as described in B. Sample with asterisk denotes that it had to be sub-cultured again as primary culture exceeded 95 % of CPE which precluded proper FACS analysis. D. Mean fluorescence intensity of positive cells detected by FACS after intracellular staining as described in B. Sample with asterisk had to be sub-cultured.

Legend:

A. CPE detection by microscopy from the cultures where clinical samples where grown. B. Percentage of positive cells detected by FACS after intracellular staining with the anti-vaccinia pAb revealed with an Alexa 488 secondary antibody from the cultures where clinical samples where grown. C. Confocal microscopy images of cells where clinical samples where grown.

## Acknowledgements

The authors acknowledge the collaboration of Gema Ballega, Pili Bonamusa, Pamela Nef, Ana Isabel Parra Manzano, Enola Crespillo, Aroa Muñoz, Núria Vaquero, Elisa Molina Molina and F. Hoffmann-La Roche Ltd. In addition, the authors thank Harvey Evans for the English revision.

## Author’s contribution

**Conceptualization:** Cristina Agustí and Jordi Casabona.

**Funding acquisition:** Jordi Casabona, Pere-Joan Cardona and Bonaventura Clotet.

**Field coordination of the study and follow-up of participants**: Héctor Martínez-Riveros.

**MPXV PCR assays:** Àgueda Hernández-Rodríguez, Cristina Casañ, Alexia París.

**MPXV genome amplification, sequencing and bioinformatics analysis**: Elisa Martró, Sandra Martínez-Puchol, Andreu Coello Pelegrin.

**Viral extraction, cell culture, titration, immunostaining and FACS analysis**: Nuria Izquierdo-Useros, Jordana Muñoz-Basagoiti, Marçal Gallemí, Daniel Perez-Zsolt, Dàlia Raïch-Regué.

**Statistical analysis:** Yesika Díaz and Lucía Alonso.

**Recruitment of participants:** Héctor Adell, Marta Villar, Rubén Mora and Luis Villegas.

**Writing – original draft:** Cristina Agustí.

**Writing – review & editing:** Elisa Martró, Núria Izquierdo, Jordi Casabona, Àgueda Hernández-Rodríguez, Cinta Folch, Ibrahim Sonmez, Héctor Martínez-Riveros, Jordana Muñoz-Basagoiti, Marçal Gallemí, Cristina Casañ.

All authors contributed to the article and approved the submitted version.

