## Supplementary figures and images for "ASYMPTOMATIC MONKEY POX VIRUS INFECTION: A SELF-SAMPLING SCREENING INTERVENTION ADRESSED TO GAY, BISEXUAL AND OTHER MEN WHO HAVE SEX WITH MEN AND TRANS WOMEN IN SPAIN"

### Supplementary Figure 1

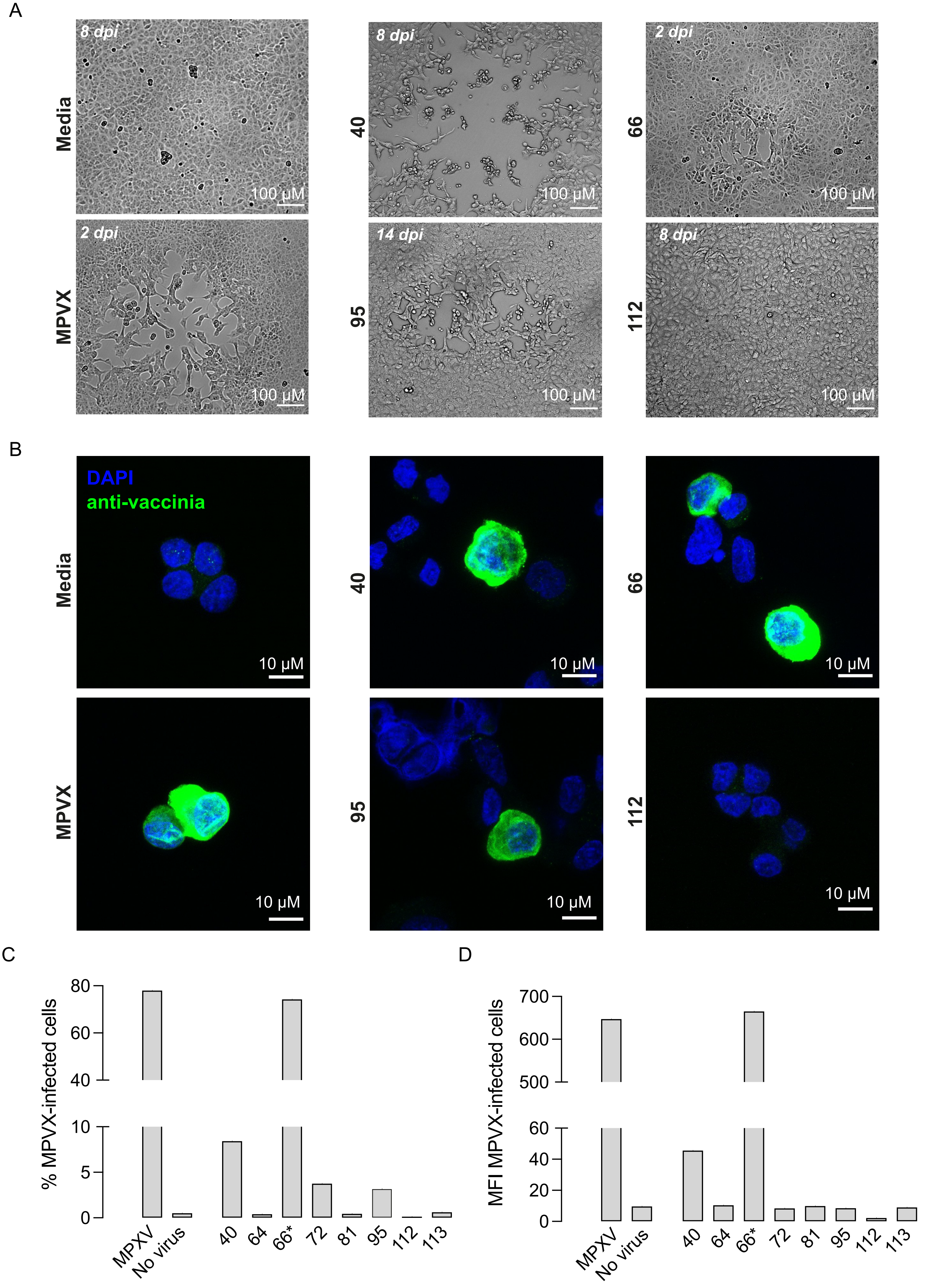


Supplementary Figure 1
